# Trends in frequency of HIV viral load and CD4 cell count monitoring among Asian cohort of adults with HIV: an analysis of the TREAT Asia HIV Observational Database, 2003-2018

**DOI:** 10.64898/2026.03.19.26348865

**Authors:** Mark Kristoffer Ungos Pasayan, Awachana Jiamsakul, Evy Yunihastuti, Iskandar Azwa, Jun Yong Choi, Nagalingeswaran Kumarasamy, Anchalee Avihingsanon, Romanee Chaiwarith, Yu-Jiun Chan, Vohith Khol, Sasisopin Kiertiburanakul, Man Po Lee, I Ketut Agus Somia, Sanjay Pujari, Cuong Duy Do, Thach Ngoc Pham, Fujie Zhang, Suwimon Khusuwan, Oon Tek Ng, Junko Tanuma, Yasmin Gani, Rohidas Borse, Jeremy Ross, Rossana Ditangco, IeDEA Asia-Pacific

**Affiliations:** Research Institute for Tropical Medicine, Muntinlupa City, Philippines; The Kirby Institute, UNSW Sydney, Sydney, Australia; Faculty of Medicine Universitas Indonesia - Dr. Cipto Mangunkusumo General Hospital, Jakarta, Indonesia; Infectious Diseases Unit, Department of Medicine, University of Malaya, Kuala Lumpur; Division of Infectious Diseases, Department of Internal Medicine, Yonsei University College of Medicine, Seoul, South Korea; CART CRS, Voluntary Health Services, Chennai, India; HIV-NAT, Thai Red Cross AIDS Research Centre, Bangkok, Thailand; Division of Infectious Diseases and Tropical Medicine, Department of Medicine, Faculty of Medicine, Chiang Mai University, Chiang Mai, Thailand; Taipei Veterans General Hospital, Taipei, Taiwan; National Center for HIV/AIDS, Dermatology & STDs, Phnom Penh, Cambodia; Faculty of Medicine Ramathibodi Hospital, Mahidol University, Bangkok, Thailand; Queen Elizabeth Hospital, Hong Kong SAR; Faculty of Medicine Udayana University & Sanglah Hospital, Bali, Indonesia; Institute of Infectious Diseases, Pune, India; Bach Mai Hospital, Hanoi, Vietnam; National Hospital for Tropical Diseases, Hanoi, Vietnam; Beijing Ditan Hospital, Capital Medical University, Beijing, China; Chiangrai Prachanukroh Hospital, Chiang Rai, Thailand; Tan Tock Seng Hospital, National Centre for Infectious Diseases, Singapore; National Center for Global Health and Medicine, Tokyo, Japan; Hospital Sungai Buloh, Sungai Buloh, Malaysia; BJ Government Medical College and Sassoon General Hospital, Pune, India; TREAT Asia, amfAR - The Foundation for AIDS Research, Bangkok, Thailand

## Abstract

**Introduction:** Viral load (VL) testing is the recommended approach for monitoring antiretroviral therapy (ART) effectiveness, while guidelines recommend targeted CD4 testing after ART initiation. This study examined trends in VL and CD4 testing frequencies, as well as the relationship with AIDS diagnosis and mortality among people with HIV in the Asia-Pacific region.

**Methods:** We included adults enrolled in the Treat Asia HIV Observational Database (TAHOD) between 2003-2018 who had been on ART for ≥1 year. VL and CD4 testing rates were analysed using Poisson regression models. Associations between testing frequency and AIDS diagnosis or mortality were evaluated using Fine and Gray competing risk regression.

**Results:** Among 8,446 patients, VL testing rates remained steady at 1 per person-year (PYS) between 2003-2018. Increased VL testing was associated with more frequent CD4 testing (>2 tests in the previous year; IRR=1.57, 95%CI 1.53-1.60), later follow-up years (2008-2012: IRR=1.15, 95%CI 1.12-1.18; 2013-2015: IRR=1.07, 95%CI 1.04-1.10), older age (31-40 years: IRR=1.06, 95%CI 1.03-1.08; 41-50 years: IRR=1.08, 95%CI 1.05-1.11; >50 years: IRR=1.07, 95%CI 1.03-1.11), higher current VL (401-1000 copies/mL: IRR=1.16, 95%CI 1.09-1.24; >1000 copies/mL: IRR=1.07, 95%CI 1.04-1.11), initial ART regimen (NRTI+PI: IRR=1.07, 95%CI 1.04-1.10; other combinations: IRR=1.11, 95%CI 1.05-1.17), and higher country income levels (upper-middle: IRR=2.17, 95%CI 2.11-2.23; high: IRR=3.14, 95%CI 3.03-3.26). CD4 testing rates decreased from 2.04 to 1.06/PYS over the same period. Lower CD4 testing frequency was associated with HIV exposure mode (MSM: IRR=0.94, 95%CI 0.92-0.96; IDU: IRR=0.93, 95%CI 0.90-0.97; other/unknown: IRR=0.90, 95%CI 0.87-0.93), higher current CD4 (201-350 cells/µL: IRR=0.95, 95%CI 0.93-0.97; 351-500 cells/µL: IRR=0.89, 95%CI 0.87-0.91; >500 cells/µL: IRR=0.85, 95%CI 0.83-0.87) and receiving an NRTI+PI first-line combination (IRR=0.96, 95% CI 0.94-0.98). VL and CD4 testing frequencies were not significantly associated with AIDS diagnosis. However, having > 2 CD4 tests in the previous year was associated with higher mortality risk.

**Conclusion:** The trends in the rates for CD4 and VL testing in the region between 2003-2018 were significantly affected by demographic, clinical and socio-economic factors. Recognizing these factors is critical to optimizing differentiated monitoring strategies and improving outcomes for PWH in the region.

## BACKGROUND

In 2016, the World Health Organization (WHO) recommended routine viral load (VL) testing as the preferred method to monitor antiretroviral treatment (ART) effectiveness and for the early detection of treatment failure. VL testing was recommended at 6 months and at 12 months after ART initiation and then annually thereafter if the patient remains clinically stable [1]. Studies have shown that mortality, loss to follow up and CD4 cell count trajectories were more favourable in care programs where VL monitoring is routinely implemented [2]. Evidence suggests that CD4 cell count monitoring has little clinical benefit among patients who are virologically suppressed. Population-based studies showed that CD4 cell count monitoring can be reduced or be made optional among patients responding well to ART [3–5]. This led to changes in guidelines recommendations for targeted rather than routine CD4 testing after ART initiation.

Despite these recommendations, the global scale-up of routine VL testing remains slow, particularly in low- and middle-income countries, mainly due to cost, limited staff training and weak laboratory systems [6]. Limited studies have assessed the effect of WHO’s recommendations on actual VL and CD4 monitoring practices during ART, and there remains uncertainty around the role of CD4 cell count monitoring, reflected in the inconsistencies among clinical guidelines and protocols [4]. Moreover, the extent to which increased global reliance on VL testing and WHO recommendations have influenced HIV treatment monitoring practices in the Asia-Pacific region remains unknown.

To determine changes in CD4 and VL testing frequencies over time in the region, we analysed data from the TREAT Asia HIV Observational Database (TAHOD), a multicentre, observational cohort study involving 21 study sites in the Asia-Pacific region.

## METHODS

### Study population

Patients enrolled in TAHOD from 2003 to 2018 who have been on ART for at least 1 year and have been enrolled in the cohort for at least 1 year were included in the study. Patients who were transferred out, became lost to follow-up (LTFU) or died within one year of cohort enrolment were excluded. Detailed methods of data collection have been published previously [7].

### CD4 and VL testing rates

CD4 and VL testing rates (per person-years, /PYS) during follow-up were plotted across calendar years between 2003 to 2018, with their 95% confidence interval (CI). A patient was considered to have been in follow-up during a given calendar year if they had a recorded clinic visit date or laboratory testing date at any time during that year. Follow-up time was censored on the date of last visit or December 31, 2018 if the patient was seen beyond 2018.

### Factors associated with CD4 and VL testing

We assessed factors associated with CD4 and VL testing rates by fitting two separate repeated measure Poisson regression models with random effect on patient. Risk time was left truncated at cohort entry or date of ART initiation, whichever occurred last, and ended on the date of last visit or December 31, 2018. In the VL analysis, each VL test during follow-up was considered the outcome of interest. CD4 testing frequency in the previous 12 months was included as a time-updated covariate to determine association between CD4 tests and current VL testing rates. Similarly, in the CD4 analysis, each CD4 measurement was counted as the outcome of interest, with VL testing frequency in the previous 12 months included as a covariate.

### AIDS diagnosis

To determine if there was a relationship between CD4 and VL testing frequency and the development of a new AIDS diagnosis, we conducted a Fine and Gray competing risk regression analysis with LTFU as competing risk. AIDS diagnosis was defined as a Centers for Disease Control and Prevention (CDC) grade C diagnosis. Risk time was left truncated at cohort entry or ART initiation and ended on the date of new AIDS diagnosis. Patients without an AIDS diagnosis and who did not become LTFU were censored on the date of last follow-up or December 31, 2018. CD4 and VL testing frequency in the previous 12 months were included as time-updated covariates.

### Survival

Survival time was analysed using Fine and Gray competing risk regression with LTFU as competing risk. Our inclusion criteria required patients to be alive and in follow-up for at least 1 year from cohort entry or ART start date. To remove this survival bias, risk time was left truncated at 1 year after cohort entry or 1 year after ART initiation and ended on the date of death. Patients who did not have at least 1 day of follow-up during this analysis period were excluded. Patients who were alive or transferred out were censored on the date of last visit. Patients with follow-up in 2019 were censored on December 31, 2018. CD4 and VL testing frequency were included as time-updated covariates.

Other covariates included calendar year of follow-up, current ART regimen, age at ART initiation, sex, HIV exposure mode, viral load, CD4 counts, initial ART regimen, hepatitis B/C coinfection, AIDS illness. World Bank country income level was adjusted a priori. All regression models were fitted using backward stepwise selection process. Covariates with p<0.10 in the univariate analyses were chosen for inclusion in the multivariate model. Covariates with p<0.05 were considered statistically significant in the final multivariate model.

Ethics approvals were obtained from the local ethics committees of all participating sites, the data management and biostatistical program (The Kirby Institute, UNSW Sydney), and the coordinating centre (TREAT Asia/amfAR). Data management and statistical analyses were performed using SAS software version 9.4 (SAS Institute Inc., Cary, NC, USA) and Stata software version 16.1 (Stata Corp., College Station, TX, USA).

## RESULTS

Among 8,446 patients included from 21 sites in 12 countries (Cambodia, China, India, Indonesia, Japan, Malaysia, Philippines, Singapore, South Korea, Taiwan, Thailand, and Vietnam), 69.6% male, and the median age at ART initiation was 34 years (interquartile range (IQR) 29-41). The main mode of HIV exposure was heterosexual contact (64.1%). The median CD4 cell count at ART initiation was 137 cells/µL (IQR 45-241) and the median VL was 83000 copies/mL (IQR 21300-250000). The median follow-up time during 2003-2018 was 8.2 years (IQR 4.4-11). Table 1 describes patient characteristics in detail.

**Table 1.** Patient characteristics.

|  | **Total patients (%)**<br>**N=8446 (100)** |
| --- | --- |
| **Year of ART initiation** |  |
| 2003-2007 | 3969 (47.0) |
| 2008-2012 | 3457 (40.9) |
| 2013-2015 | 693 (8.2) |
| 2016-2018 | 327 (3.9) |
| **Age at ART initiation (years)** | Median = 34, IQR (29-41) |
| ≤30 | 2633 (31.2) |
| 31-40 | 3579 (42.4) |
| 41-50 | 1581 (18.7) |
| >50 | 653 (7.7) |
| **Sex** |  |
| Male | 5875 (69.6) |
| Female | 2571 (30.4) |
| **Mode of HIV Exposure** |  |
| Heterosexual contact | 5410 (64.1) |
| MSM | 1857 (22.0) |
| Injecting drug use | 581 (6.9) |
| Other/unknown | 598 (7.1) |
| **Pre-ART viral load (copies/mL)** | Median = 83000, IQR (21300-250000) |
| ≤100000 | 2165 (25.6) |
| >100000 | 1764 (20.9) |
| Not tested | 4517 (53.5) |
| **Pre-ART CD4 (cells/μL)** | Median = 137, IQR (45-241) |
| ≤50 | 1928 (22.8) |
| 51-100 | 971 (11.5) |
| 101-200 | 1696 (20.1) |
| >200 | 2436 (28.8) |
| Not tested | 1415 (16.8) |
| **Initial ART Regimen** |  |
| NRTI+NNRTI | 7000 (82.9) |
| NRTI+PI | 1244 (14.7) |
| Other | 202 (2.4) |
| **Hepatitis B co-infection** |  |
| Negative | 5935 (70.3) |
| Positive | 678 (8.0) |
| Not tested | 1833 (21.7) |
| **Hepatitis C co-infection** |  |
| Negative | 5276 (62.5) |
| Positive | 870 (10.3) |
| Not tested | 2300 (27.3) |
| **Country Income Level** |  |
| Lower Middle | 3532 (41.8) |
| Upper Middle | 3112 (36.8) |
| High | 1802 (21.3) |

### VL and CD4 testing rates

Incidence rates of VL and CD4 testing, and their 95% CI are shown in Figure 1. VL testing rates remained stable at approximately 1/PYS between 2003-2018. CD4 testing rates, however, decreased from 2.04/PYS in 2003 to 1.06 in 2018. Table 2 shows that, overall, the VL testing rate was 1.12/PYS during 2003-2018.

**Figure 1:**
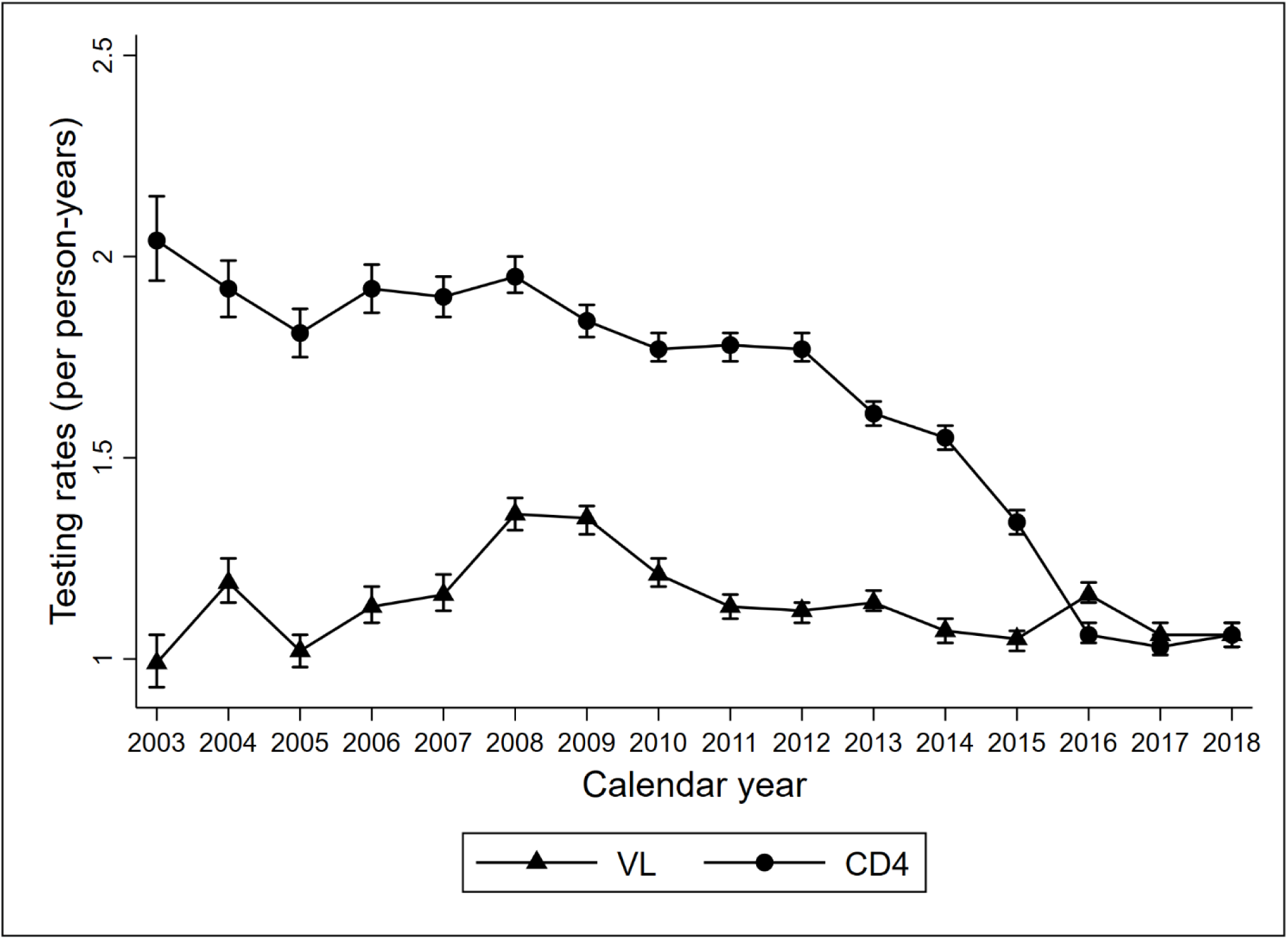
Trends in CD4 and VL testing rates over time.

**Table 2:**
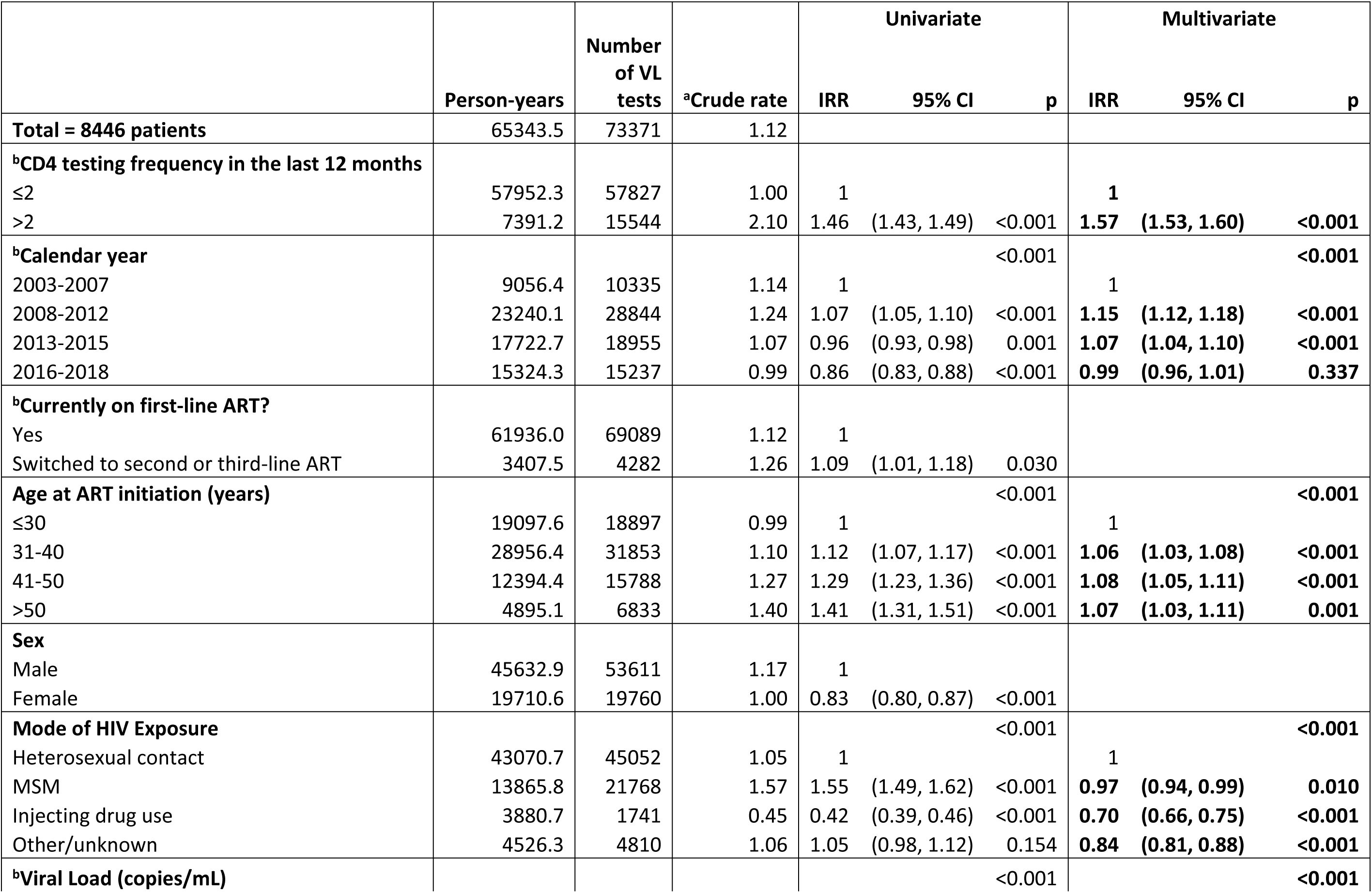
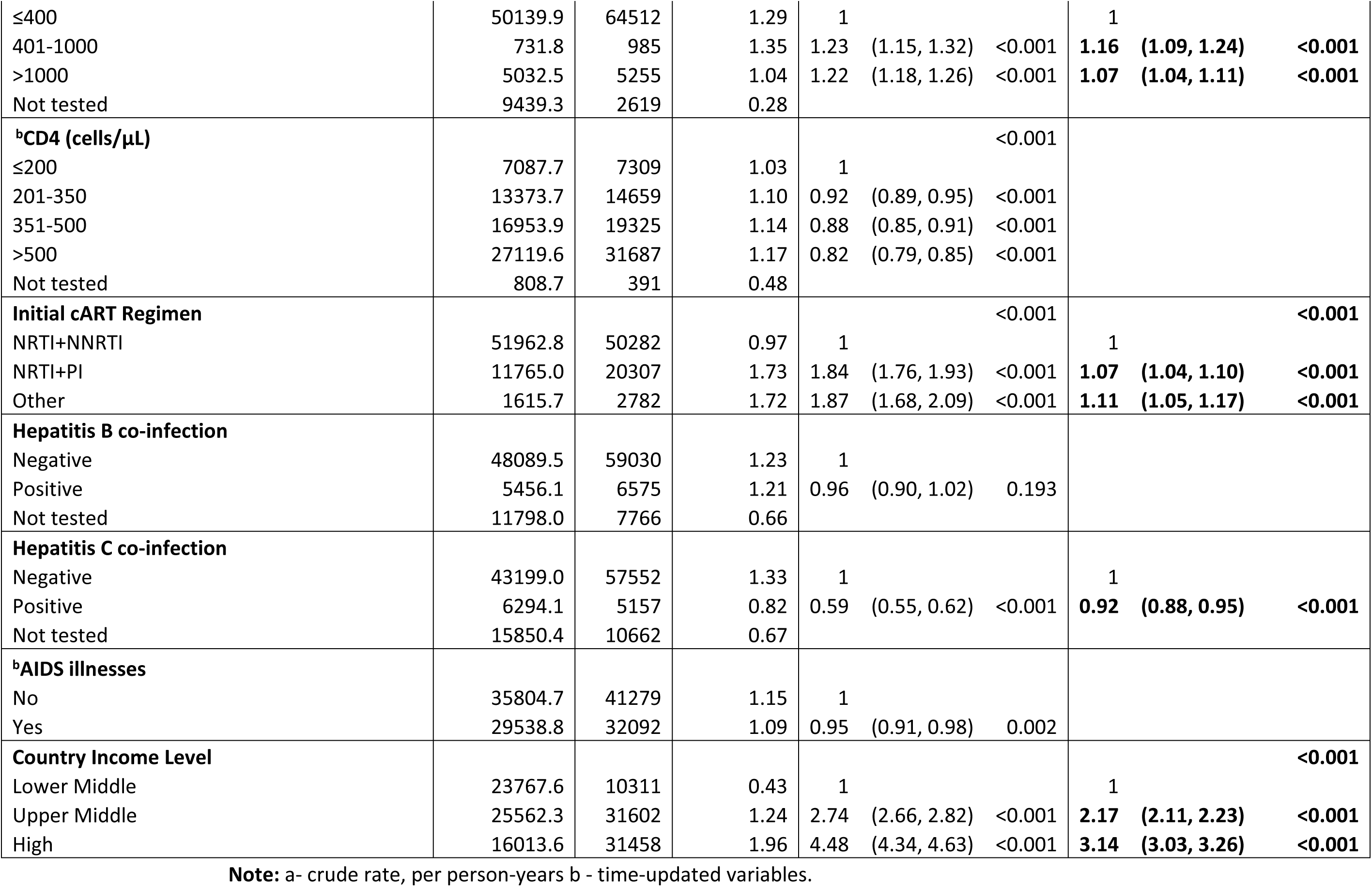
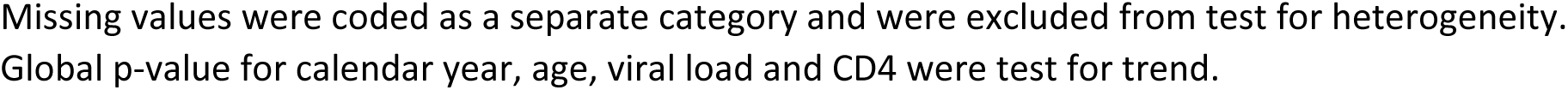
Factors associated with VL testing rates.

Having more than two CD4 tests in the previous 12 months was significantly associated with higher VL testing rates (incidence rate ratio (IRR) = 1.57, 95% CI 1.53-1.60, p < 0.001). Compared to the 2003-2007 period, follow-up in later years was associated with higher VL testing (2008-2012: IRR = 1.15, 95% CI 1.12-1.18, p < 0.001; and 2013-2015: IRR = 1.07, 95% CI 1.04-1.10, p < 0.001). Additional factors associated with higher VL testing rates were older age compared to age ≤ 30 years (31-40 years: IRR = 1.06, 95% CI 1.03-1.08, p < 0.001; 41-50 years: IRR = 1.08, 95% CI 1.05-1.11, p < 0.001; and >50 years: IRR = 1.07, 95% CI 1.03-1.11, p = 0.001), higher current VL (401-1000 copies/mL: IRR = 1.16, 95% CI 1.09-1.24, p < 0.001; and >1000 copies/mL: IRR = 1.07, 95% CI 1.04-1.11, p < 0.001), having an initial ART combination other than nucleoside reverse transcriptase inhibitor + non-nucleoside reverse transcriptase inhibitor (NRTI+NNRTI) (NRTI+ protease inhibitor (PI): IRR = 1.07, 95% CI 1.04-1.10, p < 0.001; and other combinations: IRR = 1.11, 95% CI 1.05-1.17, p < 0.001), and residence in upper or higher income countries (upper-middle: IRR = 2.17, and 3.14, respectively ; both p < 0.001). Conversely, lower VL testing rates were observed among men who have sex with men (MSM) (IRR = 0.97, 95% CI 0.94-0.99, p = 0.010; people who inject drugs (IRR = 0.70, 95% CI 0.66-0.75, p < 0.001), those with other/unknown mode of exposure (IRR = 0.84, 95% CI 0.81-0.88, p < 0.001), and being hepatitis C co-infected (IRR = 0.92, 95% CI 0.88-0.95, p < 0.001).

Overall, CD4 testing rate occurred slightly more frequently than the VL testing, at 1.55/PYS (Table 3). VL testing more than once in the past year was associated with higher CD4 testing rates (IRR=1.41, 95% CI 1.39-1.44, p<0.001). Higher CD4 testing rates were also observed among those with older ages (31-40: IRR=1.04, 95% CI 1.02-1.06, p<0.001; 41-50: IRR=1.04, 95% CI (1.02-1.06), p=0.001; and >50: IRR=1.06, 95% CI 1.02-1.09, p=0.001). Female sex was also associated with higher CD4 testing rates (IRR=1.05, 95% CI 1.03-1.07, p<0.001). Higher VL (401-1000 copies/mL: IRR=1.17, 95% CI 1.11-1.24, p<0.001; and >1000 copies/mL: IRR=1.08, 95% CI 1.05-1.11, p<0.001), receiving other combination ART regimen compared to NRTI+NNRTI (IRR=1.06, 95% CI 1.01-1.11, p=0.023), and being from countries other than lower-middle income countries (upper-middle: IRR=1.05, 95% CI 1.02-1.07, p<0.001; and high: IRR=1.40, 95% CI 1.36- 1.44, p<0.001) were also associated with higher CD4 testing rates.

**Table 3:**
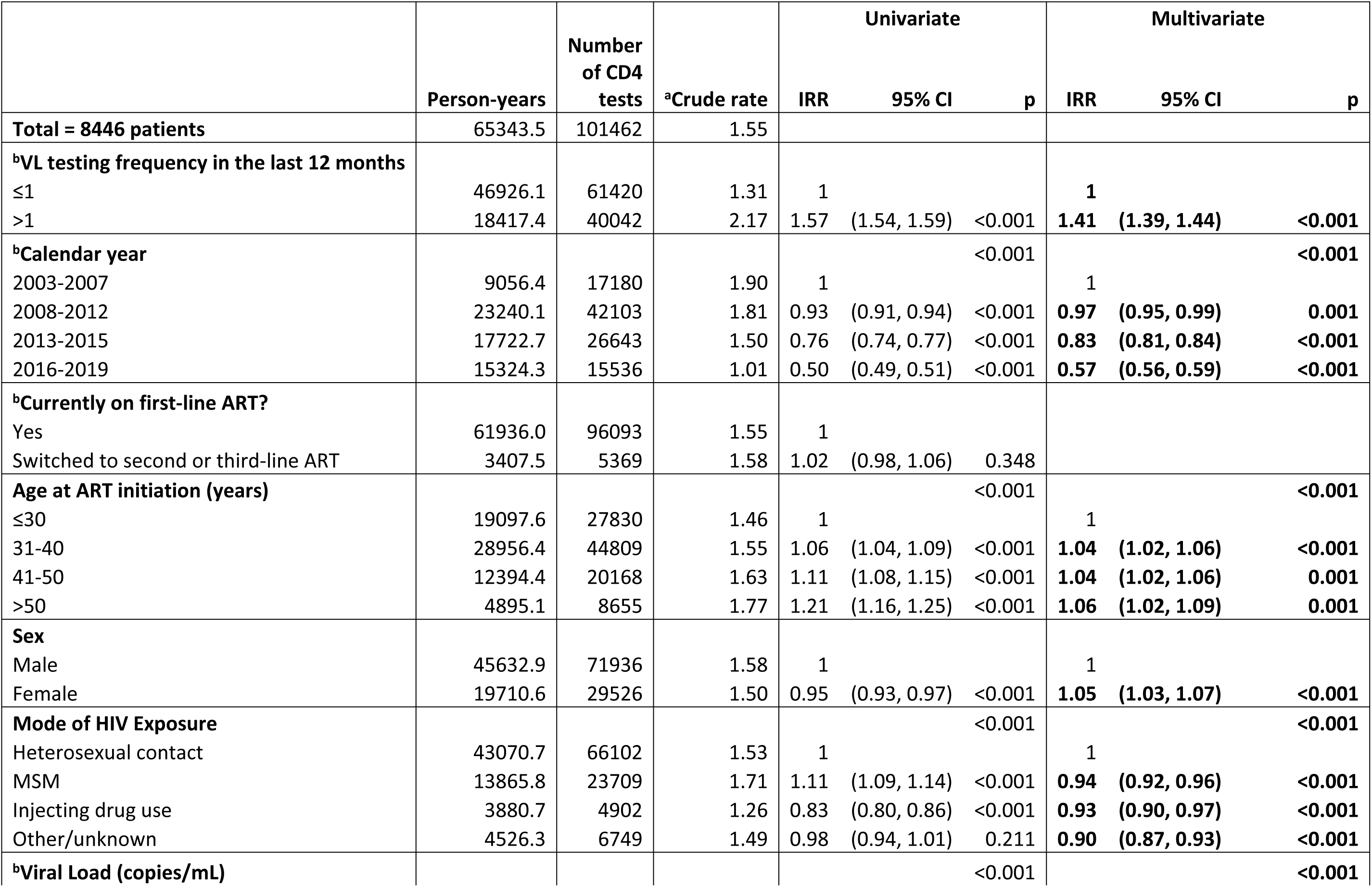
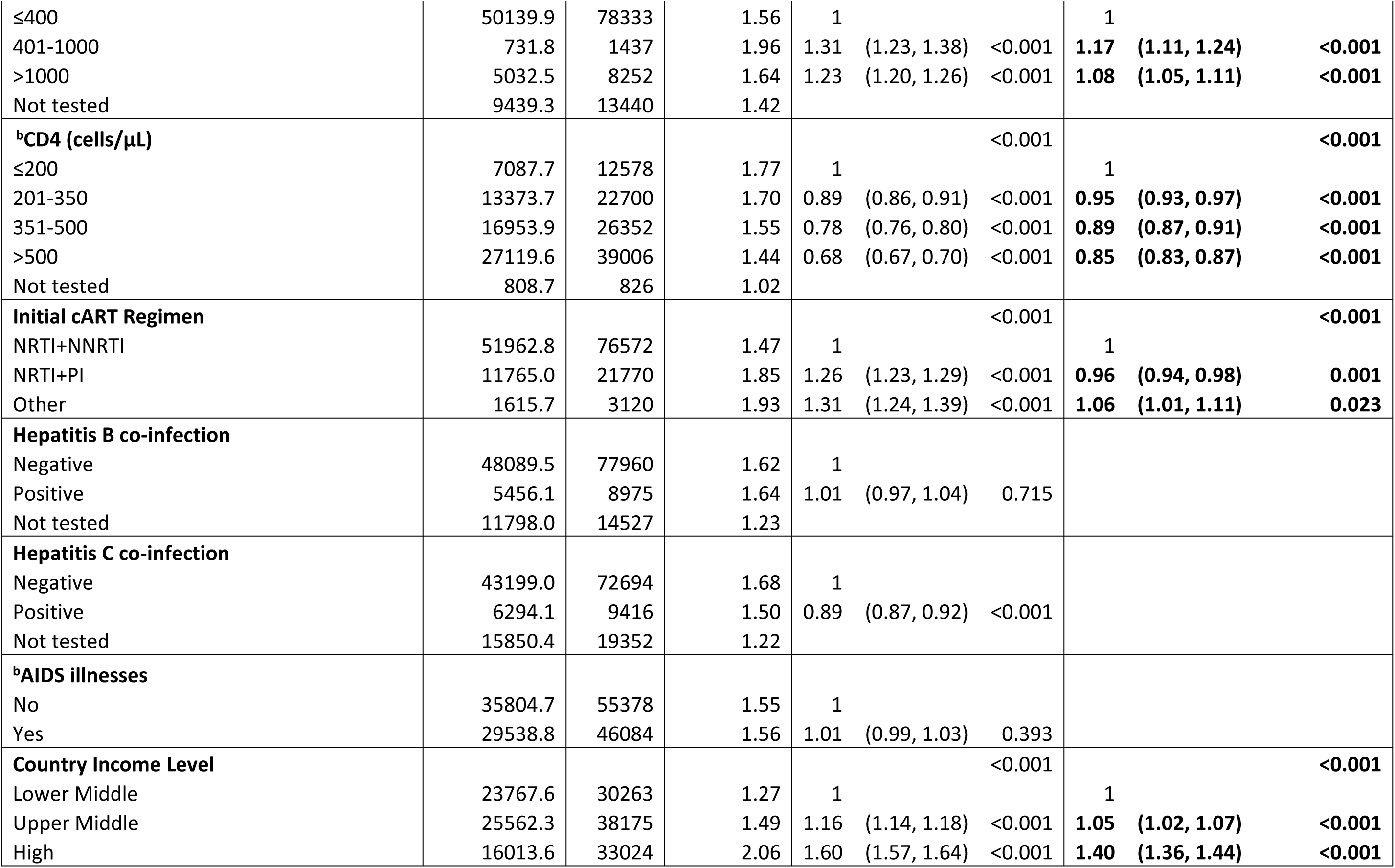
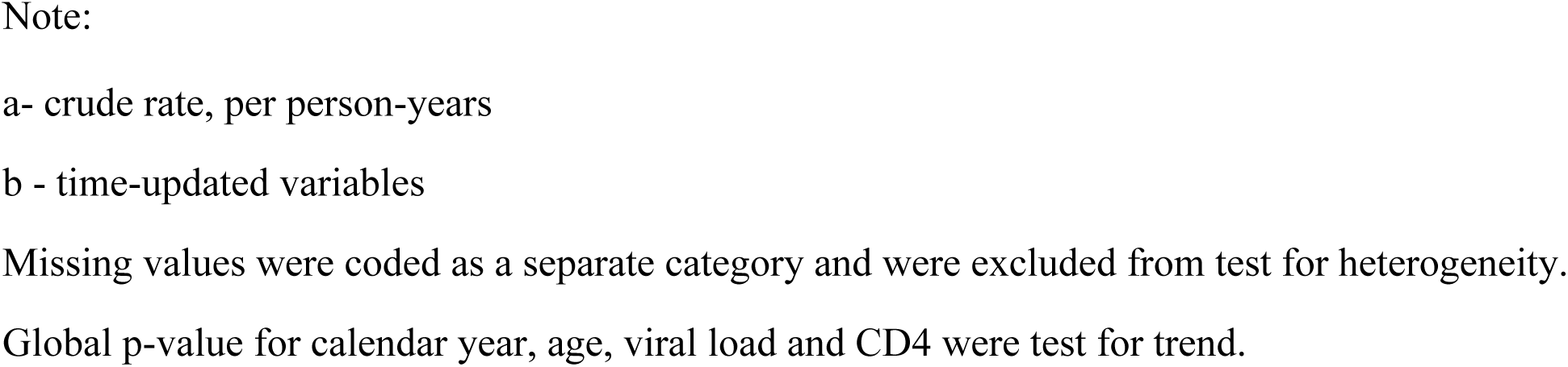
Factors associated with CD4 testing rates.

There was a trend towards decreased CD4 testing over the years (2008-2012: IRR=0.97, 95% CI 0.95-0.99, p=0.001; 2013-2015: IRR=0.83, 95% CI (0.81-0.84, p<0.001; and 2016-2019: IRR=0.57, 95% CI 0.56-0.59, p<0.001), compared to 2003-2007. Factors associated with lower CD4 testing rates were non-heterosexual mode of HIV exposure (MSM: IRR=0.94, 95% CI 0.92-0.96, p<0.001; injecting drug use: IRR=0.93, 95% CI 0.90-0.97, p<0.001; and other/unknown: IRR=0.90, 95% CI 0.87-0.93, p<0.001), higher current CD4 counts >200 cells/µL (201-350 cells/µL: IRR=0.95, 95% CI 0.93-0.97, p <0.001; 351-500 cells/µL: IRR=0.89, 95% CI 0.87-0.91, p<0.001; and >500 cells/µL: IRR=0.85, 95% CI 0.83-0.87, p<0.001), and use of NRTI+PI first-line combination compared to NRTI+NNRTI (IRR=0.96, 95% CI 0.94-0.98, p=0.001).

### AIDS diagnoses

Among 8,466 patients, 759 (9.0%) developed an AIDS-defining illness during follow-up time, with an incidence rate of 1.25/100PYS (Table 4). VL and CD4 testing frequencies were not significantly associated with AIDS diagnoses. However, a reduced risk of AIDS was observed with later calendar years (2008-2012: sub-hazard ratio (SHR)=0.65, 95% CI 0.55-0.77, p<0.001; 2013-2015: SHR=0.54, 95% CI 0.42-0.69, p<0.001; and 2016-2018: SHR=0.51, 95% CI 0.38-0.69, p<0.001) compared to 2003-2007. Higher CD4 counts >200 cells/µL (201-350 cells/µL: SHR=0.48, 95% CI 0.39-0.58, p<0.001; 351-500 cells/µL: SHR=0.29, 95% CI 0.22-0.37, p<0.001; and >500 cells/µL: SHR=0.23, 95% CI 0.17-0.29, p<0.001) were also associated with a reduced risk of AIDS. Being from upper-middle income countries was associated with a decreased risk for an AIDS-defining illness compared to lower- / middle-income countries (SHR=0.54, 95% CI 0.44-0.66, p<0.001). A higher risk for new AIDS diagnoses was seen among those who switched from first-line ART (SHR=2.45, 95% CI 1.96-3.07, p<0.001), having high VL compared to VL ≤400 copies/mL (401-1000 copies/mL: SHR=2.38, 95% CI 1.47-3.84, p<0.001; and >1000 copies/mL: SHR=2.63, 95% CI 2.12-3.27, p<0.001), and having been diagnosed with an AIDS illness prior to ART initiation (SHR=1.69, 95% CI 1.45-1.98, p<0.001).

**Table 4:**
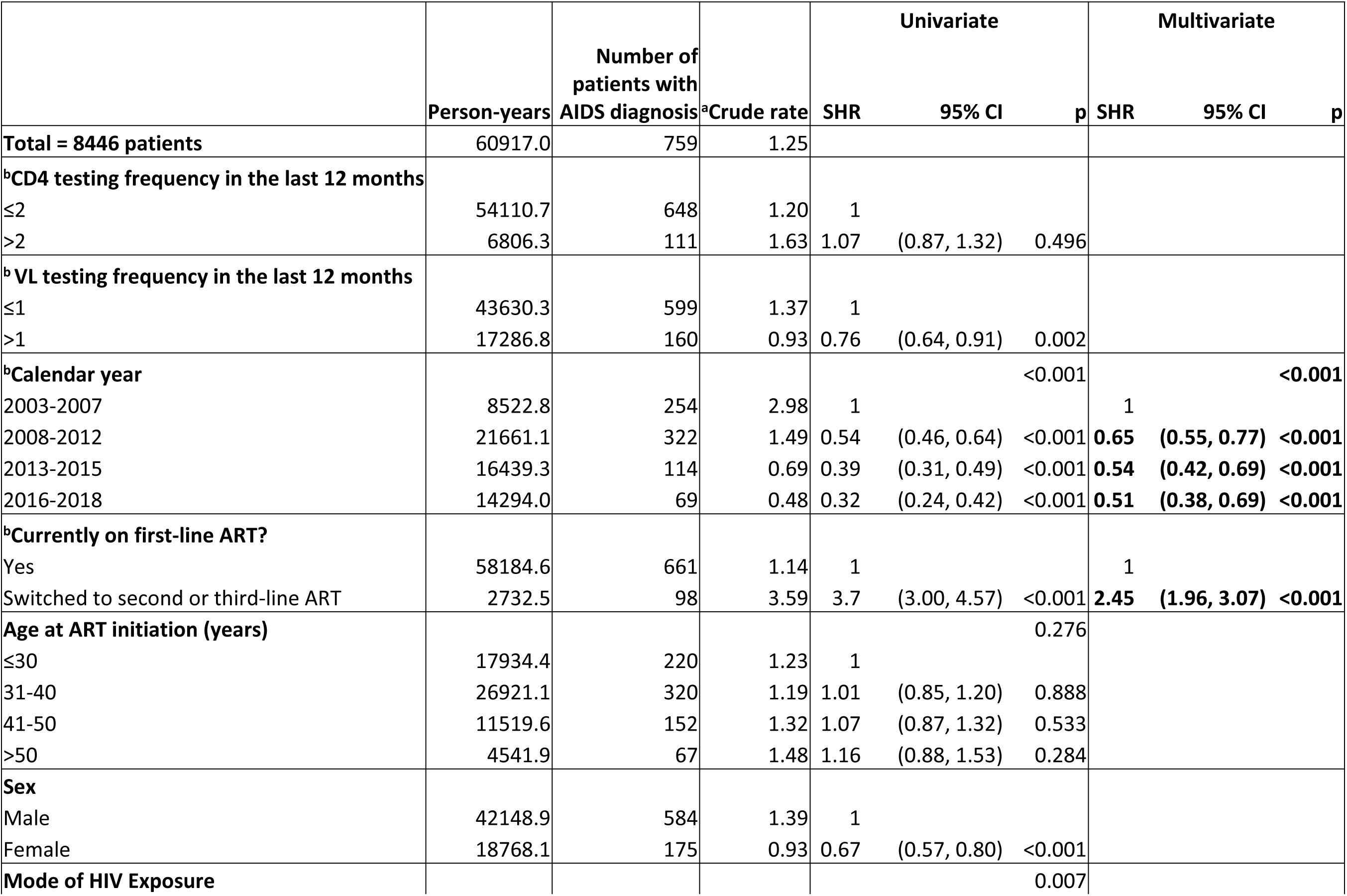
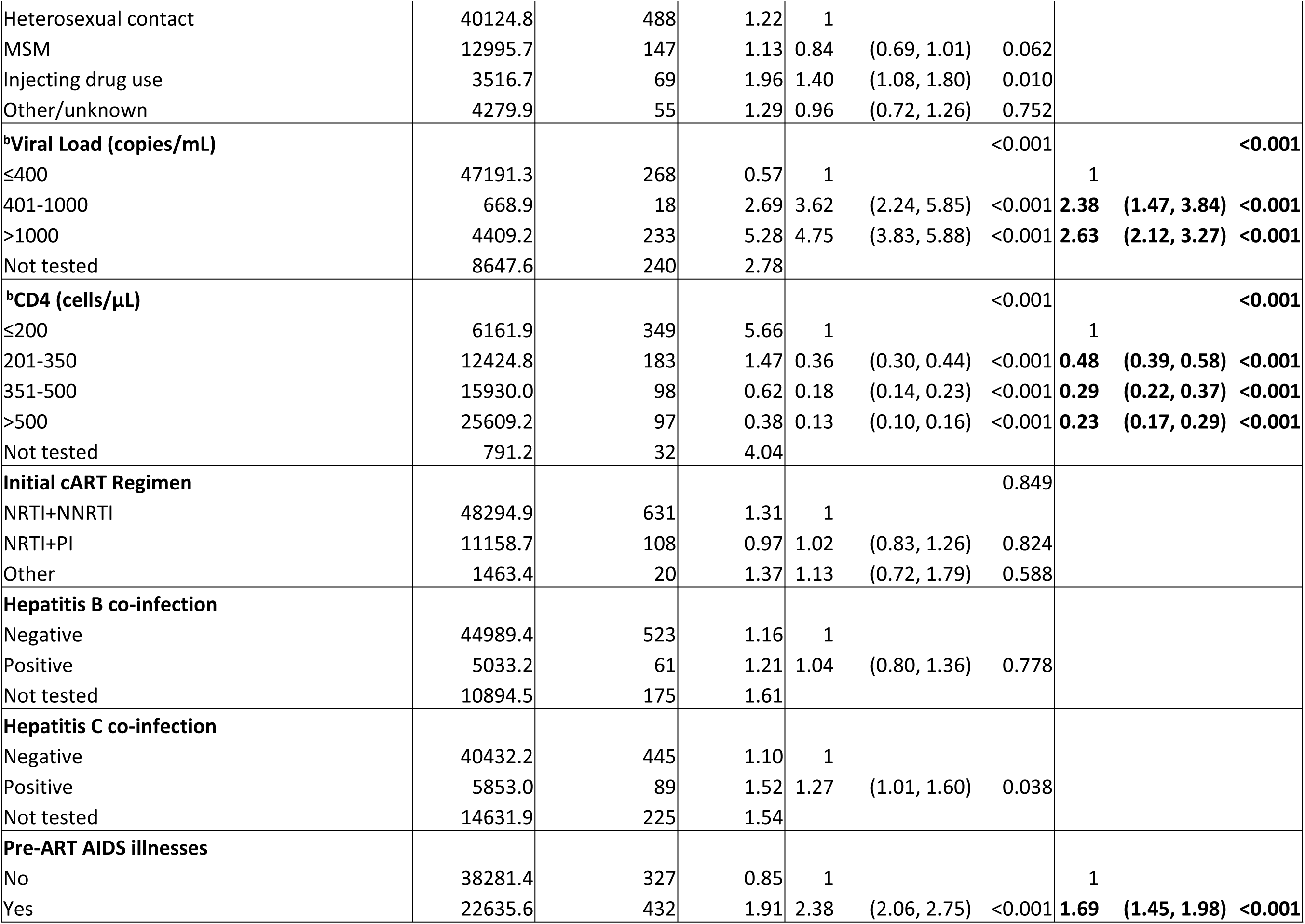
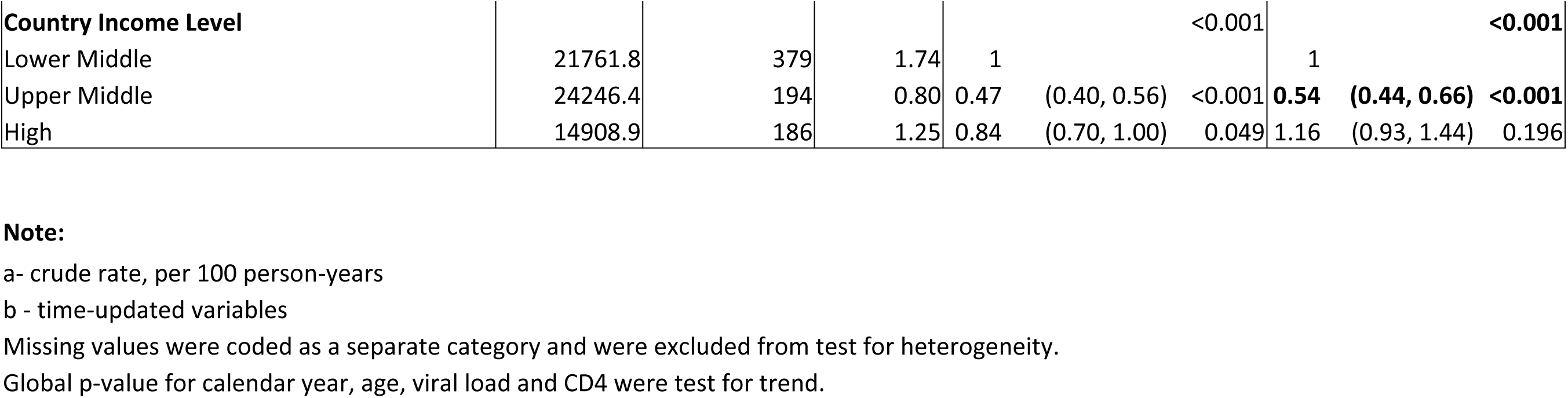
Factors associated with new AIDS diagnoses.

### Survival

Of the 8,466 patients, 8,198 (97%) had at least 1 day of follow-up during the analysis period and were included in the survival analysis. There were 357 (4.4%) deaths and 1,103 (13.5%) LTFU. The mortality rate was 0.63/100PYS (Table 5). VL testing frequency and calendar year of follow-up were not associated with survival. However, having CD4 tests >2 times in the previous year was associated with increased mortality (SHR=1.43, 95% CI 1.06-1.91, p=0.017). Other factors associated with increased risk for mortality were having switched from first-line ART (SHR=1.52, 95% CI 1.09-2.10, p=0.012), older age at ART initiation (41-50 years: SHR=1.55, 95% CI 1.13-2.11, p=0.006; and >50 years: SHR=3.56, 95% CI 2.58-4.90, p<0.001), higher VL >1,000 copies/mL (SHR=2.22, 95% CI 1.62-3.05, p<0.001), being co-infected with hepatitis B (SHR=1.57, 95% CI 1.14-2.17, p=0.006) and hepatitis C (SHR=1.56, 95% CI 1.11-2.19, p=0.011), and having a prior AIDS diagnosis (SHR=2.35, 95% CI 1.84-3.02, p<0.001). Higher CD4 counts were associated with reduced risk for mortality (201-350 cells/µL: SHR=0.38, 95% CI 0.29-0.51, p<0.001; 351-500 cells/µL: SHR=0.18, 95% CI 0.13-0.26, p<0.001; and >500 cells/µL: SHR=0.10, 95% CI 0.07-0.15, p<0.001).

**Table 5:**
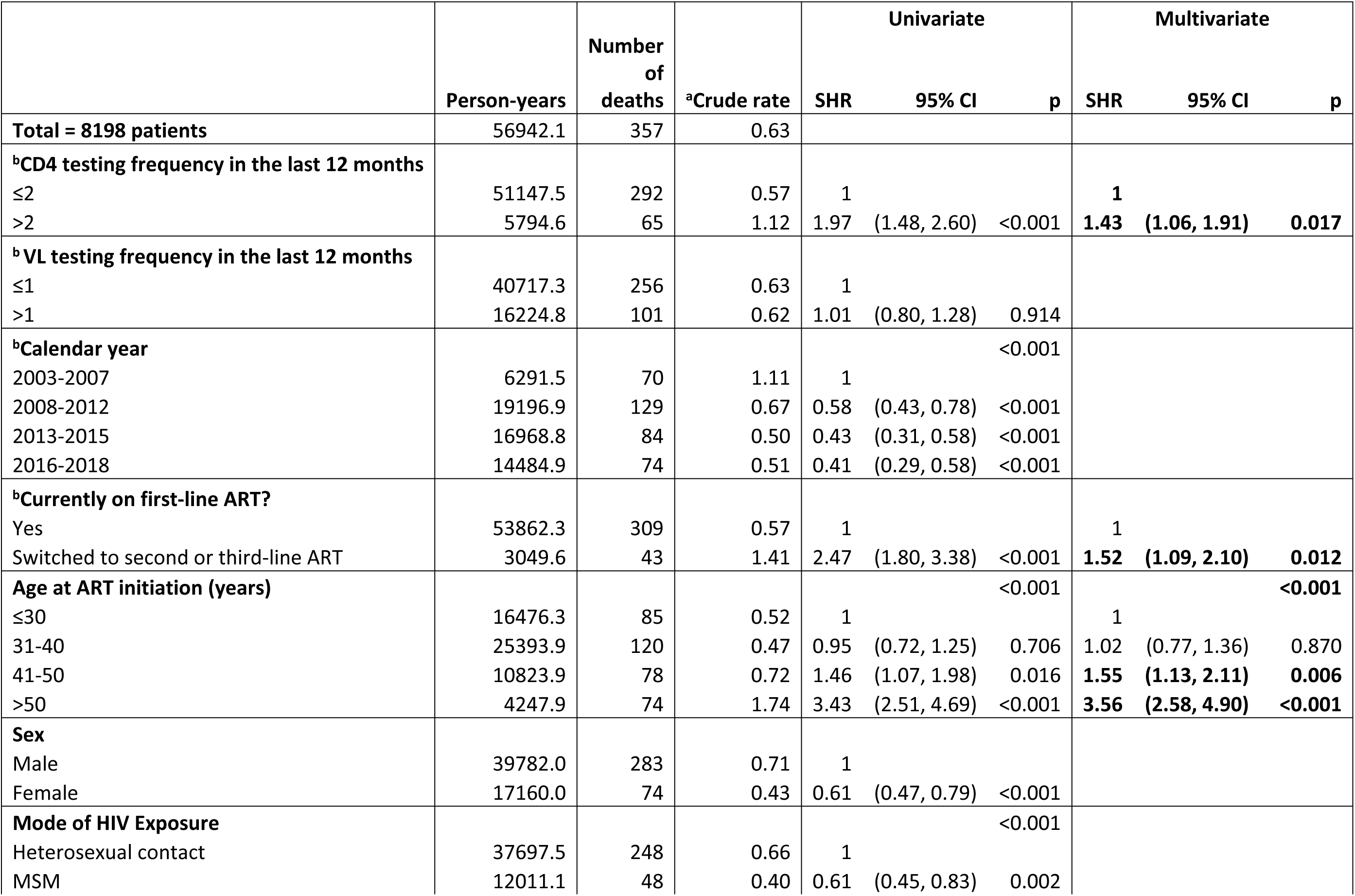
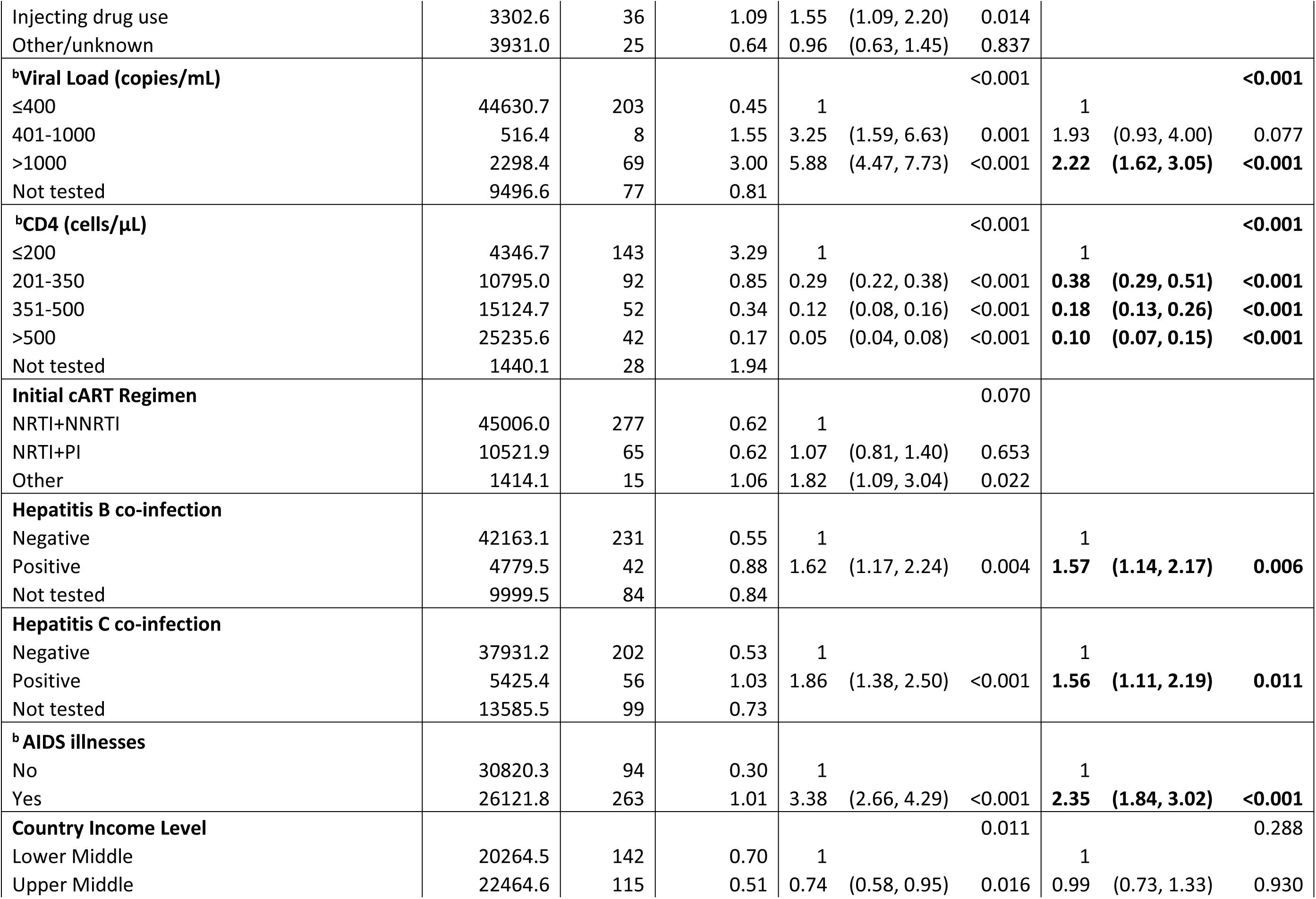
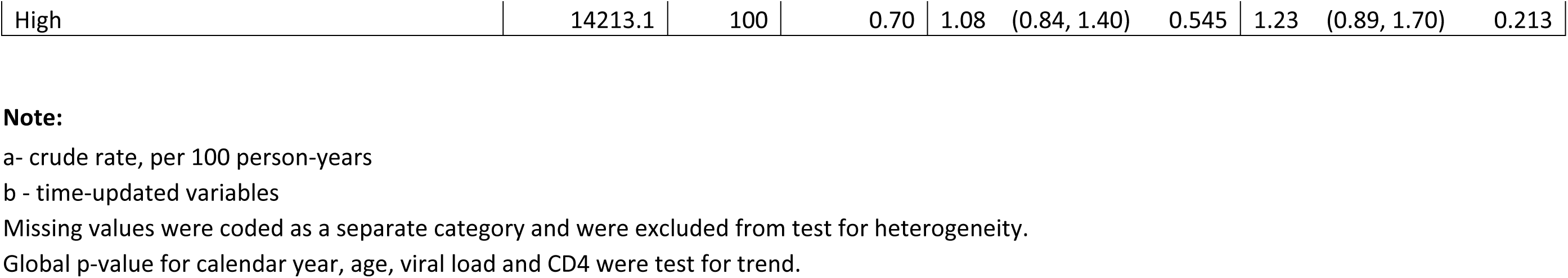
Factors associated with survival.

## DISCUSSION

Although a steady rate was seen in VL testing from 2003-2018, higher testing trends were associated with later follow-up years 2008-2015, in higher income countries, with more frequent CD4 testing, among those aged >30 years at ART initiation, with current VL >400 copies/ml, and among those receiving ART combination other than NRTI+NNRTI. These findings align with a 26-country multiregional analysis which found that VL monitoring increased significantly only in upper-middle and high-income countries following WHO’s “Treat-All” policy, with limited adoption in low- / low middle-income countries (L/LMIC) due to cost, infrastructure, and training constraints. While there was no immediate change in VL monitoring after ART initiation at Treat-All policy was adopted, trends significantly increased afterwards more frequently seen in higher income countries in this TAHOD cohort [8]. Despite the WHO guidelines outlining strategies on how to scale up VL testing, expectations on the implementation of routine VL testing and the availability of testing have not been met in L/LMIC.

The frequency of VL testing is affected by the rate of CD4 testing and vice versa since these tests are usually done during the same clinic visit. Furthermore, VL and CD4 testing rates also reflect the level of retention in care [9–11]. Our findings of higher rates of VL and CD4 testing among older patients are similar to the results of other cohort studies where a higher proportion of older adults are retained in HIV care and with better virologic outcomes compared to younger adults. Younger adults are more likely to be mobile, migrate for work, and more engaged in daily activities, which may contribute to a higher risk of loss to follow-up; hence, less frequent VL and CD4 monitoring [12–14].

The high VL and CD4 testing rates were also observed among those with detectable VL (>400 copies/ml) and among patients who were not on NRTI+NNRTI. More frequent clinic visits and optimized follow-up laboratory testing are expected from these patients with virologic failure and on second-line ART regimens. Adherence support is important to ensure retention in care [14].

Lower rates of VL testing were observed among MSM, injecting drug users, those with unknown modes of exposure, and those with hepatitis C co-infection. The low rate of VL testing seen among MSM could have been influenced by health-seeking behavior resulting in inconsistent follow-up care. This may also be attributed to the variations in the structure of HIV treatment, care, and support services for MSM. [15]. Furthermore, younger age, disease stage, and lack of access to relevant information on HIV were also found to be poor predictors of HIV care retention and subsequent VL and CD4 testing [16]. Patients with hepatitis C infection had decreased frequencies in VL monitoring, which is reflective of the care cascade drop-off among patients with HIV and HCV co-infection as described in one study [17]. These findings highlight the need to improve retention in care among younger patients, MSM, those with co-morbidities and even those with good immunologic and virologic responses.

The decreasing trends in the frequency of CD4 testing were observed after 2010 when CD4 level was one of the criteria for ART eligibility [18]. This trend continued over the years when it was recommended to start ART regardless of the CD4 at enrolment into care and became even less frequent when WHO recommended VL testing to monitor ART failure [19–20]. The questionable utility and cost-effectiveness of CD4 monitoring on how it could influence care and the changes in the ART guidelines can explain this trend [21]. Similarly, CD4 testing frequency declined while VL testing increased from 2008 to 2017 in ART programs in 6 Southern African countries. This was attributed to the implementation of the Treat-All policy and guidelines, and preference for VL for treatment monitoring, resulting in a decline in pre-ART and post-ART CD4 testing [22].

Frequencies of VL and CD4 testing were not associated with new diagnoses of AIDS-defining illnesses during the follow-up period. However, factors contributing to the decreasing trend in AIDS diagnosis remain the same as previously described [23]. This study reported the association between increased frequency of CD4 testing and increased mortality. CD4 cell count is the most prognostic factor for death in the era of effective combination ART and plays a critical role in identifying individuals with advanced HIV disease and high mortality risks. Patients with low CD4 despite being on effective ART will still need regular CD4 level monitoring, more intensive follow-up, and care [24–26].

CD4 testing remains an important component of the continuum of HIV care, from assessing disease progression to evaluating an individual’s need for opportunistic infections screening, prophylaxis, and treatment. With the release of the guidance and recommendations on the usage of CD4 monitoring, we have seen changes in the prioritization and use of these tests in relation to demographic, behavioural, clinical, and socio-economic factors. Nonetheless, CD4-based risk stratification remains essential, and its timely use of results for identifying advanced HIV disease will optimize clinical outcomes [26].

Since 2013, WHO has recommended the use of VL to monitor ART treatment response. However, the capacity of treatment facilities in Asia-Pacific to do routine VL testing remains unmet, resulting in delays in the implementation of the current recommendations. As VL testing becomes more accessible to resource-limited countries, treatment sites adjust operationally, and more data from clinical settings become available on how VL testing frequency could affect treatment outcome. Results from this test should lead to a differentiated service delivery and better clinical decision-making.

## CONCLUSION

Both CD4 and VL testing are essentials in the continuum of comprehensive HIV care. VL testing is integral for monitoring treatment efficacy and guiding ART optimization, while CD4 testing continues to play a key role in risk stratification, particularly for identifying individuals with advanced HIV disease and high mortality risk. The frequency of both tests in this Asia-Pacific cohort was significantly affected by demographic, clinical, and socio-economic factors – particularly in limited-resource settings.

Results of our study highlight the importance of differentiated strategies to improve the operational laboratory capacity of treatment sites and scale-up VL testing while maintaining the strategic use of CD4 testing for risk stratification. Eventually, the value and cost-effectiveness of doing these tests as part of HIV care delivery will be reflected in improved retention in care, reduced mortality, and sustained treatment success among people with HIV in the Asia-Pacific region.

## Data Availability

ll relevant data are within the manuscript and its Supporting Information files.

## Acknowledgements

This study is an initiative of TREAT Asia, a program of amfAR, The Foundation for AIDS Research, with support from the U.S. National Institutes of Health’s National Institute of Allergy and Infectious Diseases, the *Eunice Kennedy Shriver* National Institute of Child Health and Human Development, the National Cancer Institute, the National Institute of Mental Health, the National Institute on Drug Abuse, the National Heart, Lung, and Blood Institute, the National Institute on Alcohol Abuse and Alcoholism, the National Institute of Diabetes and Digestive and Kidney Diseases, and the Fogarty International Center, as part of the International Epidemiology Databases to Evaluate AIDS (IeDEA; U01AI069907). The Kirby Institute is funded by the Australian Government Department of Health and Ageing, and is affiliated with the Faculty of Medicine, UNSW Sydney. The content of this publication is solely the responsibility of the authors and does not necessarily represent the official views of any of the governments or institutions mentioned above.

## TAHOD study members

V Khol, V Ouk, C Pov, V Bun, National Center for HIV/AIDS, Dermatology & STDs, Phnom Penh, Cambodia; FJ Zhang, HX Zhao, N Han, Beijing Ditan Hospital, Capital Medical University, Beijing, China; MP Lee, PCK Li, TS Kwong, KY Yeung, Queen Elizabeth Hospital, Hong Kong SAR; N Kumarasamy, S Poongulali, B Faith, VHS-Infectious Diseases Medical Centre, Chennai Antiviral Research and Treatment Clinical Research Site (CART CRS), Voluntary Health Services, Chennai, India; S Pujari, K Joshi, S Gaikwad, A Chitalikar, Institute of Infectious Diseases, Pune, India; RT Borse, V Mave, I Marbaniang, S Nimkar, BJ Government Medical College and Sassoon General Hospital, Pune, India; IKA Somia, TP Merati, NM Dewi Dian Sukmawati, F Yuliana, Faculty of Medicine Udayana University - Ngoerah Hospital, Bali, Indonesia; E Yunihastuti, B Wicaksana, A Widhani, S Maria, Faculty of Medicine Universitas Indonesia - Dr. Cipto Mangunkusumo General Hospital, Jakarta, Indonesia; J Tanuma, H Gatanaga, H Uemura, Y Koizumi, National Center for Global Health and Medicine, Tokyo, Japan; JY Choi, JH Kim, JE Park, Division of Infectious Diseases, Department of Internal Medicine, Yonsei University College of Medicine, Seoul, South Korea; YM Gani, TK Heng, NA Misnan, SK Chidhambaram, Hospital Sungai Buloh, Sungai Buloh, Malaysia; I Azwa, A Kamarulzaman, SF Syed Omar, S Ponnampalavanar, University Malaya Medical Centre, Kuala Lumpur, Malaysia; RA Ditangco, MK Pasayan, JB Sornillo, Research Institute for Tropical Medicine, Muntinlupa City, Philippines; HP Chen, YJ Chan, PF Wu, Taipei Veterans General Hospital, Taipei, Taiwan; CS Wong, PL Lim, P A Kumar, Z Ferdous, National Centre for Infectious Diseases, Tan Tock Seng Hospital, Singapore; A Avihingsanon, N Hiranburana, C Wongvoranet, C Ruengpanyathip, HIV-NAT/Thai Red Cross AIDS and Infectious Diseases Research Centre, Bangkok, Thailand; S Kiertiburanakul, A Phuphuakrat, L Chumla, N Sanmeema, Faculty of Medicine Ramathibodi Hospital, Mahidol University, Bangkok, Thailand; R Chaiwarith, T Sirisanthana, J Praparattanapan, K Nuket, Faculty of Medicine and Research Institute for Health Sciences, Chiang Mai University, Chiang Mai, Thailand; S Khusuwan, P Kambua, S Pongprapass, J Limlertchareonwanit, Chiangrai Prachanukroh Hospital, Chiang Rai, Thailand; TN Pham, KV Nguyen, DTH Nguyen, DT Nguyen, National Hospital for Tropical Diseases, Hanoi, Vietnam; CD Do, AV Ngo, LT Nguyen, Bach Mai Hospital, Hanoi, Vietnam; AH Sohn, JL Ross, B Petersen, TREAT Asia, amfAR - The Foundation for AIDS Research, Bangkok, Thailand; MG Law, K Petoumenos, A Jiamsakul, D Rupasinghe, The Kirby Institute, UNSW Sydney, NSW, Australia.

